# Durability and cross-reactivity of SARS-CoV-2 mRNA vaccine in adolescent children

**DOI:** 10.1101/2022.01.05.22268617

**Authors:** Madeleine D Burns, Brittany P Boribong, Yannic C Bartsch, Maggie Loiselle, Jameson P Davis, Rosiane Lima, Andrea G Edlow, Alessio Fasano, Galit Alter, Lael M Yonker

## Abstract

Emergent SARS-CoV-2 variants and waning humoral immunity in vaccinated individuals have resulted in increased infections and hospitalizations. Children are not spared from infection nor complications of COVID-19, and the recent recommendation for boosters in individuals ages 12 years or older calls for broader understanding of the adolescent immune profile after mRNA vaccination. We tested the durability and cross-reactivity of anti-SARS-CoV-2 serologic responses over a six-month time course in vaccinated adolescents against the SARS-CoV-2 wild type and Omicron antigens. Serum from 77 adolescents showed that anti-Spike antibodies wane significantly over 6 months. After completion of a two-vaccine series, cross-reactivity against Omicron-specific receptor-binding domain (RBD) was seen. Evidence of waning mRNA-induced vaccine immunity underscores vulnerabilities in long-term pediatric protection against SARS-CoV-2 infection, while cross-reactivity highlights the additional benefits of vaccination. Characterization of adolescent immune signatures post-vaccination will inform guidance on vaccine platforms and timelines, and ultimately optimize immunoprotection of children.

## Introduction

As we entered the third year of the COVID-19 pandemic, SARS-CoV-2 infection rates surged due to highly infectious viral variants and waning population immunity. Despite full (two-dose) mRNA vaccination^1^, there are still increasing infections and hospitalizations among those individuals who have been vaccinated. Thus, boosters are now recommended for adults six months after full vaccination. More recently, there have been recommendations for adolescents (ages 12-18 years old) to obtain boosters, yet little is known of the adolescent post-vaccination immune profile.

While mortality from COVID-19 is lower in children compared to adults, over 12 million children have been diagnosed with COVID-19, leading to nearly 38,000 hospitalizations in the US^2^ and over 12,000 pediatric deaths worldwide^3^. Additionally, over 6,800 children in the US have been diagnosed with the post-infectious illness, Multisystem Inflammatory Syndrome in Children (MIS-C)^4^, and more than one in seven children experience long COVID-19^5^. Children are not spared from infection, and vaccination remains a critical strategy for preventing infection and transmission, reducing severity of disease, and limiting complications of COVID-19. Thus, we must understand and optimize vaccine responses across all pediatric age groups. Here, we quantified relative antibody responses immediately following the Pfizer-BioNTech mRNA vaccination and six months post-inoculation against the wildtype SARS-CoV-2 Spike, receptor-binding domain (RBD), and latest variant of concern (VOC), Omicron.

## Methods

Adolescent children (ages 12-17) assented, with parental consent, to participate in the MGH Pediatric COVID-19 Biorepository (MGB IRB #2020P000955)^6^. Young adults ages 18-19 years of age provided their own consent to participate. Demographic information was extracted from medical records. Blood was collected into serum separation tubes by venipuncture or by microneedle device, prior to vaccination (V0), 2-3 weeks after the initial Pfizer-BioNTech mRNA vaccination (V1), 2-4 weeks after the second dose (V2), and again 6 months after the vaccination series was complete (V6). Children were given the option to give a blood sample at any or all the time points. Blood was allowed to clot, then spun by centrifuge and serum was collected.

Serological analyses were performed using an in-house enzyme-linked immunosorbent assay (ELISA) that detects IgG against the wildtype SARS-CoV-2 Spike, the wild type RBD, or the Omicron SARS-CoV-2 VOC RBD by using the previously described method^7^. Briefly, 96-well plates were coated with 1 µg/ml of Spike or RBD overnight at 4°C in bi-carbonate buffer. The plates were then washed and serum samples were added at a 1:500 (Spike) dilution or 1:200 (RBD) dilution in duplicate for 1h at room temperature, washed and then detected with a secondary anti-IgG (Bethyl Laboratories). The secondary was washed away after 1h, and the colorimetric detector was added (TMB; Thermo Fisher) for 5 mins. The reaction was then stopped and the absorbance was acquired at 450/570nm on SpectraMax plate reader. To convert raw OD values into concentration, a two-fold dilution curve (starting at 29.8 international units) of the WHO standard (NIBSC code: 20/136) was included onto every ELISA plate. The sample concentration was interpolated from the resulting standard curve, as previously described^7^. Antibody responses at each time point were analyzed relative to the average V0 (pre-vaccination) antibody response.

Analysis was completed by Prism 9.3 using one-way ANOVA for multiple comparisons and t-test for two-way comparisons. Correlations were completed using Pearson correlation.

## Results

Seventy-seven children were enrolled in our study, with a median age of 14 years; sixty-eight children were between the ages of 12-15 years; nine were between ages of 16-19 years. Sex was equally distributed, and 19% of the population was Hispanic (**Supplemental Table 1**). Ninety-five percent (n = 73) of participants denied SARS-CoV-2 infection prior to enrollment and throughout the course of the study. Thirty-one percent (n = 24) of the children provided blood samples at all four separate time points: prior to vaccination (V0), 2-3 weeks after first vaccine dose (V1), 2-4 weeks after the second vaccine dose (V2), and 6 months after the second vaccine dose (V6).

Relative antibody responses to SARS-CoV-2, wildtype, Spike (**Figure 1A**), wildtype RBD (**Figure 1B**), and Omicron RBD (**Figure 1C**) were analyzed for each time point. Robust generation of anti-wild type and anti-Omicron antibodies were seen following the second dose of the vaccine, as compared to pre-vaccination levels (*p* < 0.0001, *p* < 0.0001, *p* < 0.0001; wild type Spike, wild type RBD, and Omicron RBD, respectively). Interestingly, there was no increase in antibodies against Omicron RBD after the first vaccine dose but a significant increase in titers was seen following a second vaccine dose. However, there was a subsequent loss of all anti-SARS-CoV-2 antibody responses by six months, as compared to the V2 time point (*p* < 0.0001, *p* < 0.0001, *p* < 0.0001; wild type Spike, wild type RBD, and Omicron RBD). By six months, antibody responses decreased to levels comparable to titers seen at the V1 time point, following the first vaccine dose. Twenty-four adolescents provided blood samples at all four time points; individual responses align with trends seen in the larger cohort (**Figure S1**). This loss of antibody titers highlights a potential vulnerability of adolescents and young adults to breakthrough infections six months after the completion of a two-dose vaccine series.

**Figure 1:**
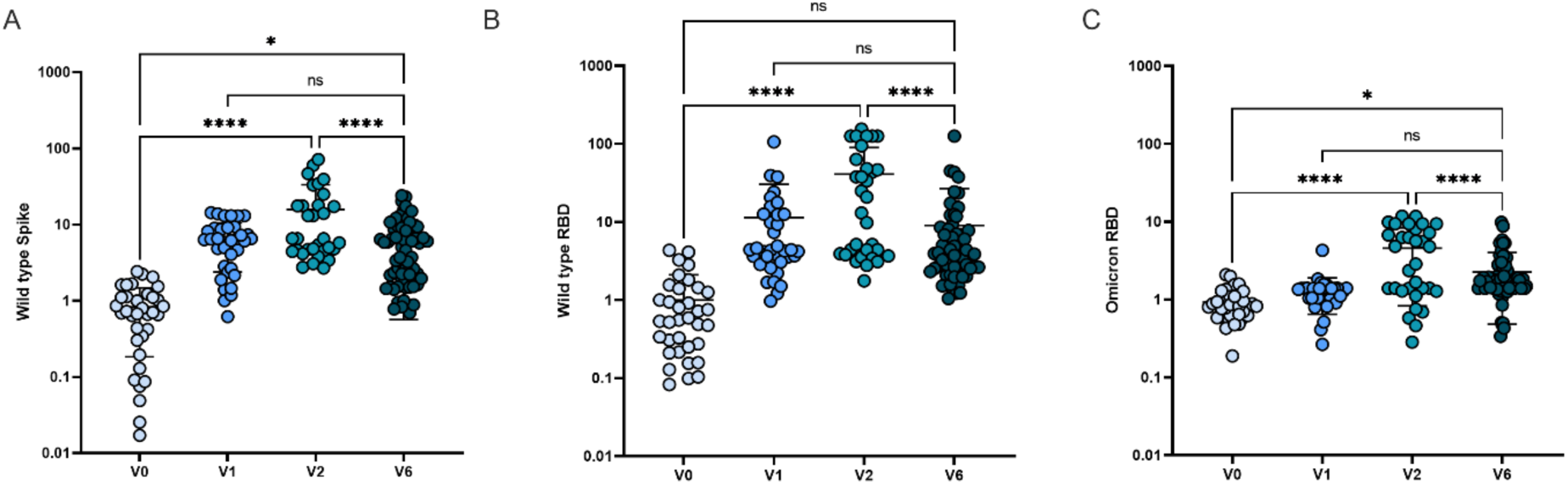
Adolescent anti-SARS-CoV-2 antibody responses over time. Relative humoral responses to a) Wild type Spike b) Wild type Receptor Binding Domain (RBD), and c) Omicron RBD are quantified prior to vaccination, 2-3 weeks following the first vaccine dose, 2-4 weeks following the second mRNA vaccine dose, and 6 months following the second mRNA vaccine dose. V0 = pre-vaccination, V1 = 2-3 weeks following the first vaccine dose, V2 = 2-4 weeks following the second mRNA vaccine dose, and V6 = 6 months following the second mRNA vaccine dose. Displayed as fold increase from baseline. Analysis by ANOVA. ns = not significant, * P < 0.05, **** P < 0.0001

As Omicron has become the predominant SARS-CoV-2 variant globally, we assessed the relationship between anti-wild type RBD and anti-Omicron RBD titers to determine if COVID-19 mRNA vaccination displayed cross-coverage and protection against the Omicron-specific SARS-CoV-2 RBD. While a single vaccination provides no protection against Omicron (**Figure 1C**), the second vaccination establishes cross-reactivity of RBD responses, although the titers remain significantly lower for Omicron RBD than for wild type RBD (**Figure 2A**; P < 0.001). At peak immunity following the second vaccination, there was a slight correlation in wild type Spike and RBD for both wild type and Omicron (**Figure 2B**, wild type *p* = 0.027, and Omicron *p* = 0.013), though this correlation plateaued at peak RBD levels. At six-months post mRNA vaccine, despite the fact that wild type RDB titers remained higher than Omicron RBD titers (**Figure 2C**; *p* < 0.01), there was a strong correlation in declining anti-wild type and Omicron RBD titers with anti-Spike titers (Figure 2D; wild type *p* = 0.0006, and Omicron *p* = 0.0007). These data underscore the need for long-term vaccine-induced pediatric immunoprotection amidst episodic surges of SARS-CoV-2 infections.

**Figure 2:**
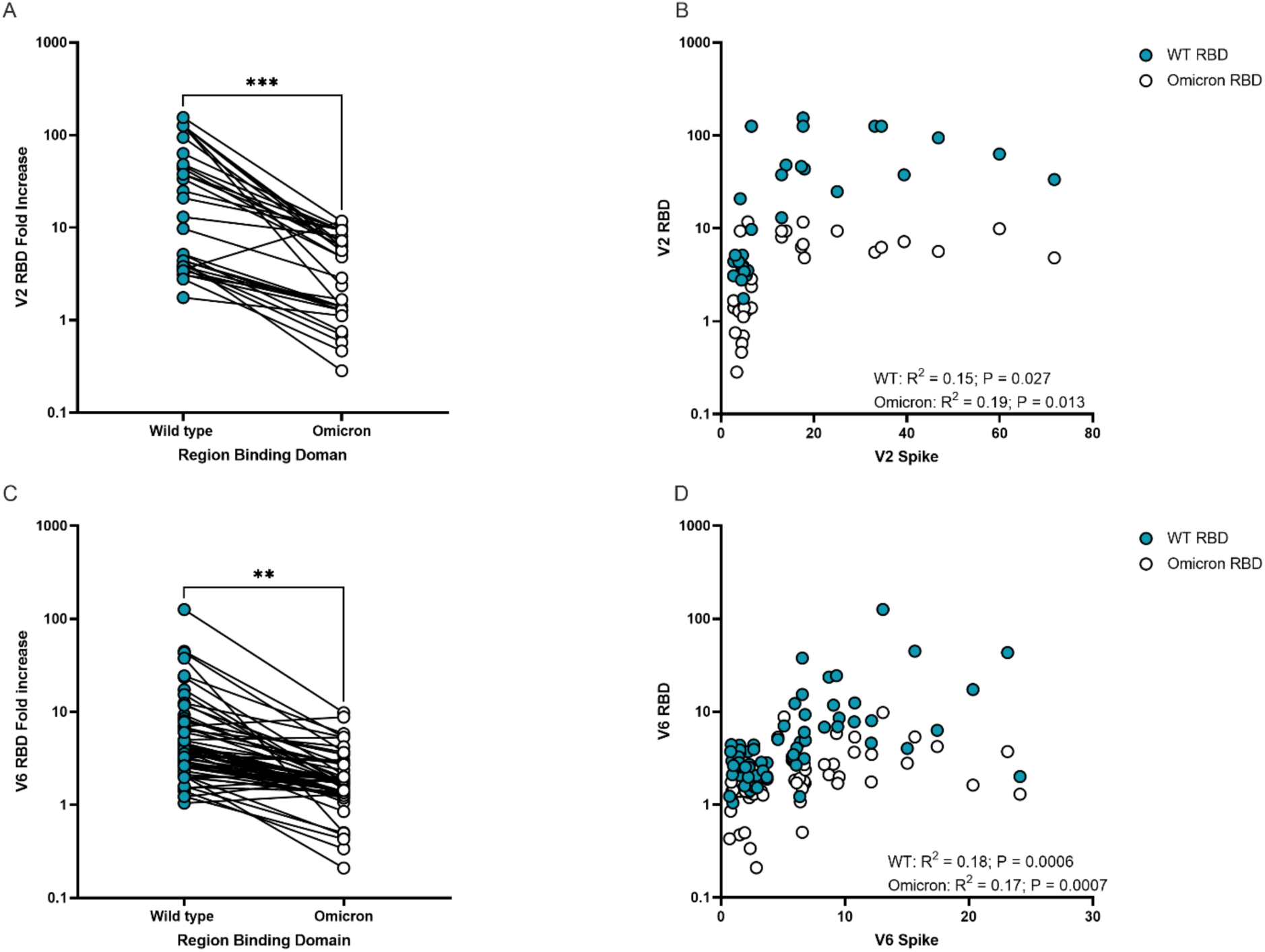
Comparison of humoral response to SARS-CoV-2 wild type and Omicron Receptor Binding Domain (RBD). A) Following the second mRNA vaccine dose, anti-RBD responses titers are compared between wild type and Omicron, B) and correlations between RBD for each variant and Spike were assessed. C) Anti-RBD titers were also compared at the 6-month time point, and correlation between RBD and Spike was again assessed. Paired analysis with t-test, correlation with Pearson correlation. WT = wild type. ** P < 0.01, *** P < 0.001

## Discussion

As the COVID-19 mRNA vaccines represent a new vaccination platform, the longevity of immune responses needs to be characterized across all age ranges, especially in light of emerging variants. Here, we specifically focus on humoral immune responses in adolescent children against SARS-CoV-2, including responses against the highly infectious and the current predominant variant, Omicron.

As expected, and as seen in adult populations, mRNA vaccine-induced immunity wanes significantly over a 6-month time period in adolescent children. This finding demonstrates a current vulnerability for infection in adolescent children, many of whom have now received their vaccine series over six months ago. Booster vaccinations will likely play a key role in generating sustained immunity in this adolescent age group.

Encouragingly, the immune responses that have been generated display sufficient cross-coverage of the VOC, Omicron, with comparable immune responses to those reported in adults^8^. This could suggest that children develop a similar adaptive humoral immune response as seen in adults^9^, which may provide benefits to children as additional variants of concern emerge. While it is plausible that vaccine-induced anamnestic immunity may provide some level of protection in this population upon exposure, the durability of vaccine-induced immunity wanes in adolescents, and boosting may promote a robust barrier of immunity that will contribute to public health efforts to limit spread and prevent future hospitalizations and severe disease.

In conclusion, adolescent children exhibit waning antibody immune responses six months post-mRNA vaccination. mRNA boosters will play a critical role in sustaining durable immune responses in adolescent children, while also reducing pediatric infection, severe illness, and transmission as we traverse the surges of the COVID-19 pandemic.

## Data Availability

All data produced in the present study are available upon reasonable request to the authors

---

**Table S1:** Demographics of adolescents enrolled and timing of assessments for serologic responses to COVID-19 mRNA vaccination.

| **Patient Characteristics** | **Cohort (N=77)** |
| --- | --- |
| **Age at Enrollment, mean (SD)** | 14 (1.8) |
| **Male, number (%)** | 41 (53) |
| **Hispanic, number (%)** | 15 (19) |
| **Race, number (%)** |  |
| **White** | 53 (69) |
| **Black** | 2 (3) |
| **Asian** | 1 (1) |
| **Other** | 16 (21) |
| **Unknown** | 5 (6) |
| **Time Since Dose 1 and V1 Draw, average days (SD)** | 18 (3) |
| **Time Since Dose 2 and V2 Draw, average days (SD)** | 18 (5) |
| **Time Since Dose 2 and V6 Draw, average months (SD)** | 6.7 (0.5) |

**Figure S1:**
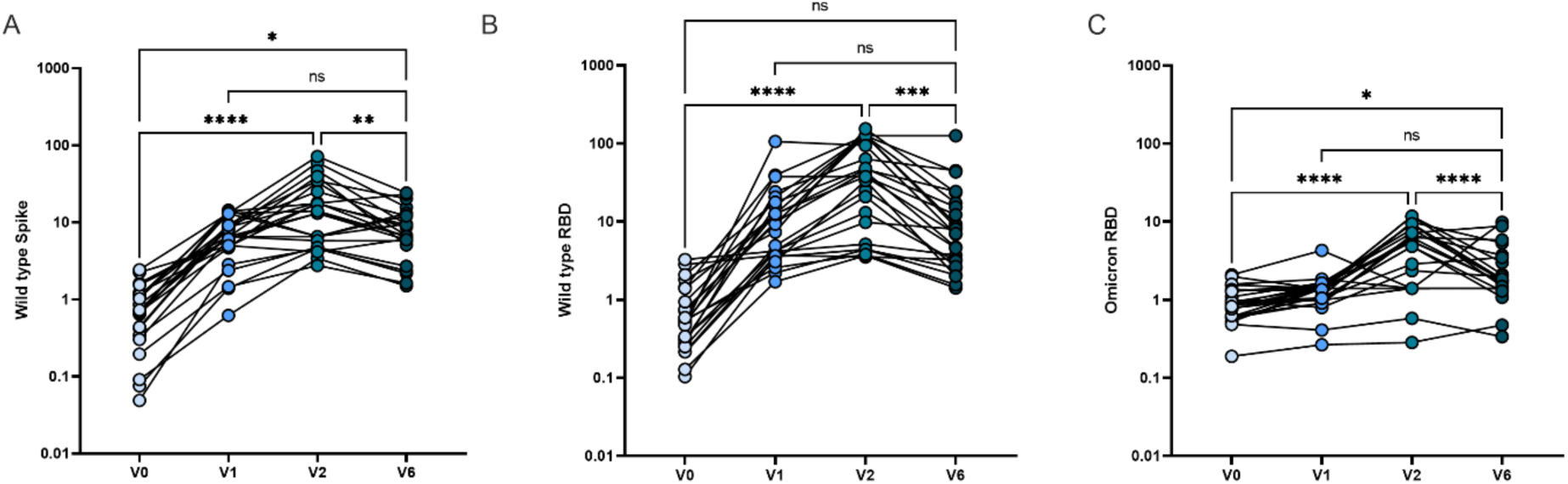
Individual anti-SARS-CoV-2 antibody responses over time. Twenty-four adolescents provided blood samples at each of the four time points. Relative humoral responses to a) Wild type Spike b) Wild type Receptor Binding Domain (RBD), and c) Omicron RBD were quantified prior to vaccination. V0 = pre-vaccination, V1 = 2-3 weeks following the first vaccine dose, V2 = 2-4 weeks following the second mRNA vaccine dose, and V6 = 6 months following the second mRNA vaccine dose. Displayed as fold increase from baseline. Analysis by ANOVA. ns = not significant, * *p* < 0.05, ** *p* < 0.01, *** *p* < 0.001 **** *p* < 0.0001

## Funding and Acknowledgements

We thank the children and families who participated in this research. We thank Nancy Zimmerman, Mark and Lisa Schwartz, an anonymous donor (financial support), Terry and Susan Ragon, and the SAMANA Kay MGH Research Scholars award for their support. We acknowledge support from Massachusetts General Hospital for Children, the Ragon Institute of MGH, MIT, and Harvard, the Massachusetts Consortium on Pathogen Readiness (MassCPR), the Musk foundation and the March of Dimes. Massachusetts Consortium on Pathogen Readiness (MassCPR), the NIH (3R37AI080289-11S1, R01AI146785, U19AI42790-01, U19AI135995-02, 1U01CA260476-01, CIVIC75N93019C00052, 5K08HL143183, R01HD100022-02S2) and the Gates Foundation Global Health Vaccine Accelerator Platform funding (OPP1146996 and INV-001650).

## Author Contributions

LMY and MDB oversaw sample collection and metadata documentation. BPB, ML, JPD and RL processed samples and performed experimental assays, BPB completed data analysis. MDB, BPB, YCB, GA and LMY analyzed and interpreted the data. AGE, AF, GA, LMY provided resources and intellectual contributions. MDB, BPB and LMY drafted the initial manuscript. All authors critically reviewed the manuscript.

## Institutional Review Board Statement

The study was conducted according to the guidelines of the Declaration of Helsinki, and was approved by the Institutional Review Board of Massachusetts General Hospital (IRB #2020P000955, approved 4/2/20).

## Informed Consent Statement

Informed consent and assent, where applicable, was obtained from all subjects involved in the study.

